# Sleep regularity outweighs sleep duration as a predictor of disease

**DOI:** 10.64898/2026.06.15.26355648

**Authors:** Daniel P. Windred, Angus C. Burns, Amy Reynolds, Kelly Sansom, Bastien Lechat, Hannah Scott, Robert Adams, Dorothee Steven, Richa Saxena, Martin K. Rutter, Frank Scheer, Sean W. Cain, Andrew J. K. Phillips

## Abstract

Sleep regularity, the consistency of sleep-wake timing from one day to the next, is more strongly associated with longevity than adequate sleep duration. Whether this relationship persists across common diseases is unknown. We compared sleep regularity *vs.* sleep duration as risk factors for 199 diseases and disorders, using ten million hours of objective sleep-wake data (N=60,998, age[mean±SD]=62.8±7.8, 55% female). Multivariable-adjusted risks of incident diseases/disorders for regular/irregular and short/adequate sleepers were compared across 9.5 years of follow-up. Irregular sleep predicted risks for 131 diseases/disorders, more than double the number predicted by short sleep duration (63). Irregular sleep was a superior predictor than short sleep duration for 90 diseases/disorders, including circulatory, metabolic, digestive, renal, infectious, neurological, and musculoskeletal conditions, and mental disorders, whereas short sleep duration was the superior predictor for only 9 diseases/disorders. For models where short sleep duration explained disease risks, 83% were improved by adding sleep regularity. Sleep regularity was a stronger predictor of diseases/disorders than sleep duration in this cohort and should be considered an essential dimension of sleep health.

## Introduction

Maintaining sleep of adequate duration is important for longevity and avoiding adverse health outcomes.^1–8^ Health guidelines for sleep typically focus on the importance of sleep duration,^9^ but are increasingly recognizing the importance of sleep regularity,^10^ the consistency of sleep-wake patterns from one day to the next.^11^ Emerging evidence indicates that sleep regularity is a robust predictor of health and wellbeing,^12–17^ and that sleep regularity may be a stronger predictor than sleep duration for some health outcomes.^12,13^ However, the comparative effects of sleep duration *vs*. sleep regularity as predictors of disease and disorder are not well understood.

Sleep duration and sleep regularity likely contribute to health status via distinct mechanisms. Insufficient sleep duration impedes important restorative functions that occur during sleep.^18,19^ Irregular sleep patterns, on the other hand, contribute to disruption of circadian rhythms,^20^ due to irregular exposure to daily time cues (‘zeitgebers’) for the circadian system, such as light exposure and meal timing. Circadian clocks are present in virtually all tissues, playing a pervasive role in most areas of human physiology.^21^ Circadian disruption and insufficient sleep have each been shown to adversely affect health, across cardiovascular, metabolic, and immune function.^22–26^

Sleep duration and regularity are largely independent dimensions of sleep behavior, with studies across a range of populations finding these two quantities are not correlated or only weakly correlated.^13^ Strategies that aim to improve sleep health in one dimension may not improve the other. Identifying the distinct contributions of sleep duration and sleep regularity to each area of health is therefore essential for developing sleep health advice, assessing risk for specific health outcomes at the individual level, and designing effective interventions.

We compared sleep duration *vs*. sleep regularity as prospective predictors of risk of 199 incident disease and disorder phenotypes, using accelerometer data and National Health Service records collected in over 60,000 UK Biobank participants. Sleep regularity was assessed as the day-to-day consistency in clock timing of sleep patterns, using a widely adopted metric developed by our research group, the Sleep Regularity Index (SRI).^11,12,27^

## Methods

### Overview

Approximately 502,000 participants were recruited to the UK Biobank between 2006 and 2010.^28^ Of these, 103,669 participants wore Axivity AX3 actigraphy devices (Axivity, Newcastle upon Tyne, UK) for up to one week between 2013 and 2016. Devices were worn on participants’ dominant wrist under naturalistic conditions and logged acceleration data at 100Hz. Detailed information on data collection is available on the UK Biobank website, as described previously.^12^ Incident diseases and disorders recorded up to November 2022 were included. This study was granted ethical approval by the North West Multi-centre Research Ethics Committee and adheres to the Strengthening the Reporting of Observational Studies in Epidemiology (STROBE) guidelines.

### Exposure: Sleep regularity and sleep duration

Methods for extraction of sleep regularity and sleep duration from actigraphy devices in this cohort has been previously reported.^12,27^ In short, sleep-wake states across each participant’s seven-day recording were determined using a validated algorithm.^29,30^ Daily sleep durations were calculated using sustained inactivity duration between algorithm-defined sleep ‘onset’ and ‘offset’ times. Daily sleep durations were averaged across recording days for each participant. Sleep Regularity Index (SRI) scores were calculated from epoch-by-epoch sleep-wake data across seven-day recordings. Each 30 sec epoch was classified as either ‘sleep’ or ‘wake’, and the SRI captured the concordance in sleep-wake state for all epoch pairs separated by 24 hours. Our methodology for SRI calculation accounted for naps, fragmented sleep, and large periods of wake during daily main sleep episodes and is accessible as an open-source R package.^27^ Out of 103,669 participants who wore actigraphy devices, 60,998 participants were included in sleep regularity and duration analyses after excluding epochs of non-wear or data corruption, days with miscalculated sleep onset or offset timing, and requiring at least five days of usable sleep data, as reported previously.^12,27^

### Outcomes: Incident Disease/Disorder Phenotypes

Disease and disorder phenotypes were derived from UK Biobank’s first occurrence data. This approach converted a large number of ICD-9 and ICD-10 diagnostic codes into a smaller number of clinically meaningful phenotypes.^31,32^ ICD diagnoses were derived from hospital, primary care, self-report, and death register records, by the UK Biobank team. Phenotype mapping data were downloaded from the Github page of the Harvard University “Translational Data Science Center for a Learning Health System” (CELEHS). Broadly defined ‘parent’ phenotypes (e.g., phe411 Ischemic Heart Disease) were used in all analyses: sub-phenotypes (e.g., phe411.4 Coronary atherosclerosis) were not analysed. We included phenotypes under the following categories: ‘Infectious diseases’, ‘Neoplasms’, ‘Endocrine/metabolic’, ‘Hematopoietic’, ‘Neurological’, ‘Mental disorders’, ‘Musculoskeletal’, ‘Sense organs’, ‘Circulatory system’, ‘Respiratory’, ‘Digestive’, ‘Genitourinary’, and ‘Dermatologic’. Phenotypes under ‘Other’, ‘Congenital’ and ‘Injuries and poisonings’ were excluded, since this work focused primarily on chronic health outcomes. Outcome categories were limited to those that affected both men and women in main analyses, and sub-group analyses for male and female groups included sex-specific outcomes. Outcomes with fewer than 240 cases across the observation period were excluded to prevent overfitting and biased estimates, by attaining a recommended >10 events-per-variable.^33^ An interactive visualization tool detailing ICD codes within each phenotype is available: https://phenomics.va.ornl.gov/phecodemap/.

### Covariates

Age, sex, ethnicity, education level, employment status, yearly household income, material deprivation (Townsend Deprivation Index),^34^ smoking status (current, previous, or never), alcohol consumption (days per week), healthy diet score,^35^ and urbanicity of residential location were derived from questionnaire data provided by participants during a baseline assessment (2006-2010). Physical activity was defined as average device acceleration across participants’ one-week activity recording.^36^ Covariates map to those used in our prior work, where detailed descriptions were provided.^37^

### Statistical Analysis

Risks of each disease/disorder phenotype were assessed with Cox proportional hazards models, including Sleep Regularity Index scores and/or mean weekly sleep duration as exposure variables (‘survival’ package in R 4.5.0). Sleep regularity and duration variables were split into categories to account for potential non-linear responses (e.g., a u-shaped response for sleep duration). Sleep Regularity Index scores were split into 0 to 30^th^, 31^st^ to 40^th^, 41^st^ to 50^th^, 51^st^ to 60^th^, 61^st^ to 70^th^, and 71^st^ to 100^th^ percentile groups. Sleep durations were split into 0 to 30^th^, 31^st^ to 40^th^, 41^st^ to 50^th^, 51^st^ to 60^th^, 61^st^ to 90^th^, and 91^st^ to 100^th^ percentile groups. These groupings were applied to compare ‘irregular’ (0-30^th^ pc.; SRI < 75.6) vs. ‘regular’ (71-100^th^ pc.; SRI ≥ 85.2) sleepers, and participants with ‘short’ (0-30^th^ pc; < 6.3 h) vs. ‘adequate’ (61-90^th^ pc.; 7.1 – 7.9 h) sleep duration (aligning with the recommended 7-8 h sleep duration for older adults^38^). Follow-up commenced one year after actigraphy (to reduce the potential influence of reverse causality), and ended at disease/disorder occurrence, mortality, or administrative endpoint of November 2022. For each outcome-specific analysis, we excluded individuals with the outcome of interest prior to the start of follow-up. Four hierarchical levels of model adjustment were applied: Model 1 was adjusted for age, sex, and ethnicity; Model 2 was further adjusted for socio-economic status (household income, education level, employment status, and material deprivation); Model 3 was further adjusted for physical activity; and Model 4 was further adjusted for healthy diet score, urbanicity, smoking status, and alcohol consumption. Models 1 and 2 contained covariates that were potentially confounding, while Models 3 and 4 contained covariates that were potentially confounding and mediating. Complete case analysis was used due to low levels of missing data. All p-values were adjusted for multiple testing using False Discovery Rate (FDR<.05). Equivalent sleep regularity and sleep duration models were compared using Akaike Information Criterion, to identify which model minimized information loss. Hierarchical models including (i) sleep regularity and (ii) sleep regularity adjusted for sleep duration were compared using Likelihood Ratio Tests to assess whether sleep duration explained additional disease/disorder risk beyond that explained by sleep regularity only (similarly, sleep duration vs. sleep-regularity-adjusted sleep duration models were compared). Sleep regularity and duration are only weakly associated in this cohort;^12^ therefore, multi-collinearity does not pose a significant concern. Weighted means of hazard ratios for each category (e.g., Circulatory system) were calculated using prevalence of each disease/disorder between activity tracking and endpoints as weights.

## Results

The analysis sample consisted of 60,998 individuals of mean age (±SD) = 62.8 (7.8), and 55% female. Descriptive statistics across demographic, socioeconomic, and lifestyle factors are provided in Table 1, for the overall analysis sample and split according to sleep regularity and sleep duration percentile groups. Missing data did not exceed 2.8% for any covariate. Observation period from one year after activity tracking to the administrative endpoint of November 2022 was mean (±SD) = 7.1 (±0.6) years.

**Table 1.** Descriptive statistics across demographic, socioeconomic, and lifestyle factors, split by sleep regularity and sleep duration percentile groups.

|  | Overall | Sleep Regularity Index percentiles |  |  |  |  |  | Sleep duration percentiles |  |  |  |  |  |
| --- | --- | --- | --- | --- | --- | --- | --- | --- | --- | --- | --- | --- | --- |
|  | N = 60,998<sup>1</sup> | 0-30<sup>th</sup><br>N = 18,309<sup>1</sup> | 31-40<sup>th</sup><br>N = 6,110<sup>1</sup> | 41-50<sup>th</sup><br>N = 6,083<sup>1</sup> | 51-60<sup>th</sup><br>N = 6,100<sup>1</sup> | 61-70<sup>th</sup><br>N = 6,096<sup>1</sup> | 71-100<sup>th</sup><br>N = 18,300<sup>1</sup> | 0-30<sup>th</sup><br>N = 18,300<sup>1</sup> | 31-40<sup>th</sup><br>N = 6,099<sup>1</sup> | 41-50<sup>th</sup><br>N = 6,100<sup>1</sup> | 51-60<sup>th</sup><br>N = 6,100<sup>1</sup> | 61-90<sup>th</sup><br>N = 18,299<sup>1</sup> | 91-100<sup>th</sup><br>N = 6,100<sup>1</sup> |
| Age (years) | 62.78 (7.79);<br>43.50 to 79.00 | 63.09 (7.81);<br>43.76 to 79.00 | 62.44 (7.82);<br>43.59 to 77.85 | 62.29 (7.78);<br>44.29 to 78.84 | 62.40 (7.83);<br>43.69 to 78.19 | 62.52 (7.76);<br>43.50 to 78.20 | 62.96 (7.73);<br>44.10 to 78.88 | 62.90 (7.86);<br>43.92 to 79.00 | 62.18 (7.91);<br>43.50 to 78.88 | 62.30 (7.81);<br>43.69 to 77.45 | 62.19 (7.75);<br>43.89 to 78.84 | 62.93 (7.71);<br>43.90 to 78.61 | 63.64 (7.56);<br>44.10 to 78.20 |
| Sex |  |  |  |  |  |  |  |  |  |  |  |  |  |
| Female | 33,621 (55%) | 9,358 (51%) | 3,350 (55%) | 3,428 (56%) | 3,434 (56%) | 3,475 (57%) | 10,576 (58%) | 8,270 (45%) | 3,299 (54%) | 3,474 (57%) | 3,510 (58%) | 11,191 (61%) | 3,877 (64%) |
| Male | 27,373 (45%) | 8,940 (49%) | 2,746 (45%) | 2,671 (44%) | 2,649 (44%) | 2,635 (43%) | 7,732 (42%) | 10,028 (55%) | 2,800 (46%) | 2,625 (43%) | 2,590 (42%) | 7,108 (39%) | 2,222 (36%) |
| Ethnicity |  |  |  |  |  |  |  |  |  |  |  |  |  |
| White | 59,067 (97%) | 17,438 (96%) | 5,866 (97%) | 5,930 (97%) | 5,893 (97%) | 5,967 (98%) | 17,973 (98%) | 17,350 (95%) | 5,894 (97%) | 5,911 (97%) | 5,964 (98%) | 17,937 (98%) | 6,011 (99%) |
| Mixed | 334 (0.5%) | 130 (0.7%) | 39 (0.6%) | 32 (0.5%) | 35 (0.6%) | 24 (0.4%) | 74 (0.4%) | 139 (0.8%) | 33 (0.5%) | 42 (0.7%) | 20 (0.3%) | 85 (0.5%) | 15 (0.2%) |
| Asian | 530 (0.9%) | 223 (1.2%) | 63 (1.0%) | 49 (0.8%) | 54 (0.9%) | 45 (0.7%) | 96 (0.5%) | 264 (1.4%) | 59 (1.0%) | 45 (0.7%) | 42 (0.7%) | 96 (0.5%) | 24 (0.4%) |
| Black | 428 (0.7%) | 234 (1.3%) | 51 (0.8%) | 31 (0.5%) | 45 (0.7%) | 24 (0.4%) | 43 (0.2%) | 272 (1.5%) | 40 (0.7%) | 41 (0.7%) | 24 (0.4%) | 41 (0.2%) | 10 (0.2%) |
| Chinese | 121 (0.2%) | 52 (0.3%) | 17 (0.3%) | 15 (0.2%) | 7 (0.1%) | 8 (0.1%) | 22 (0.1%) | 59 (0.3%) | 8 (0.1%) | 11 (0.2%) | 9 (0.1%) | 28 (0.2%) | 6 (<0.1%) |
| Other | 301 (0.5%) | 132 (0.7%) | 37 (0.6%) | 31 (0.5%) | 33 (0.5%) | 20 (0.3%) | 48 (0.3%) | 128 (0.7%) | 34 (0.6%) | 34 (0.6%) | 30 (0.5%) | 56 (0.3%) | 19 (0.3%) |
| Employment |  |  |  |  |  |  |  |  |  |  |  |  |  |
| Employed | 36,853 (61%) | 10,762 (59%) | 3,833 (63%) | 3,820 (63%) | 3,808 (63%) | 3,793 (63%) | 10,837 (59%) | 11,238 (62%) | 3,857 (64%) | 3,881 (64%) | 3,845 (63%) | 10,675 (59%) | 3,357 (55%) |
| Unemployed | 23,743 (39%) | 7,383 (41%) | 2,228 (37%) | 2,241 (37%) | 2,242 (37%) | 2,270 (37%) | 7,379 (41%) | 6,928 (38%) | 2,204 (36%) | 2,181 (36%) | 2,217 (37%) | 7,511 (41%) | 2,702 (45%) |
| Household yearly income (£) |  |  |  |  |  |  |  |  |  |  |  |  |  |
| <18k | 7,909 (13%) | 3,073 (17%) | 804 (13%) | 767 (13%) | 707 (12%) | 637 (10%) | 1,921 (11%) | 2,607 (14%) | 729 (12%) | 748 (12%) | 697 (11%) | 2,272 (13%) | 856 (14%) |
| 18-29.9k | 13,380 (22%) | 4,236 (23%) | 1,347 (22%) | 1,310 (22%) | 1,313 (22%) | 1,263 (21%) | 3,911 (21%) | 4,046 (22%) | 1,296 (21%) | 1,248 (21%) | 1,263 (21%) | 4,035 (22%) | 1,492 (25%) |

|  | Overall | Sleep Regularity Index percentiles |  |  |  |  |  | Sleep duration percentiles |  |  |  |  |  |
| --- | --- | --- | --- | --- | --- | --- | --- | --- | --- | --- | --- | --- | --- |
|  | N = 60,998<sup>1</sup> | 0-30<sup>th</sup><br>N = 18,309<sup>1</sup> | 31-40<sup>th</sup><br>N = 6,110<sup>1</sup> | 41-50<sup>th</sup><br>N = 6,083<sup>1</sup> | 51-60<sup>th</sup><br>N = 6,100<sup>1</sup> | 61-70<sup>th</sup><br>N = 6,096<sup>1</sup> | 71-100<sup>th</sup><br>N = 18,300<sup>1</sup> | 0-30<sup>th</sup><br>N = 18,300<sup>1</sup> | 31-40<sup>th</sup><br>N = 6,099<sup>1</sup> | 41-50<sup>th</sup><br>N = 6,100<sup>1</sup> | 51-60<sup>th</sup><br>N = 6,100<sup>1</sup> | 61-90<sup>th</sup><br>N = 18,299<sup>1</sup> | 91-100<sup>th</sup><br>N = 6,100<sup>1</sup> |
| 30-51.9k | 15,780 (26%) | 4,573 (25%) | 1,600 (26%) | 1,574 (26%) | 1,588 (26%) | 1,637 (27%) | 4,808 (26%) | 4,661 (26%) | 1,585 (26%) | 1,622 (27%) | 1,642 (27%) | 4,737 (26%) | 1,533 (25%) |
| 52-100k | 13,692 (23%) | 3,575 (20%) | 1,345 (22%) | 1,459 (24%) | 1,380 (23%) | 1,506 (25%) | 4,427 (24%) | 4,021 (22%) | 1,431 (24%) | 1,429 (24%) | 1,500 (25%) | 4,124 (23%) | 1,187 (20%) |
| >100k | 3,861 (6.4%) | 926 (5.1%) | 378 (6.2%) | 393 (6.5%) | 457 (7.6%) | 424 (7.0%) | 1,283 (7.0%) | 1,172 (6.5%) | 437 (7.2%) | 444 (7.3%) | 384 (6.3%) | 1,114 (6.1%) | 310 (5.1%) |
| Unknown | 5,943 (9.8%) | 1,758 (9.7%) | 575 (9.5%) | 555 (9.2%) | 594 (9.8%) | 609 (10%) | 1,852 (10%) | 1,640 (9.0%) | 578 (9.5%) | 569 (9.4%) | 585 (9.6%) | 1,883 (10%) | 688 (11%) |
| Townsend deprivation index | -1.79 (2.77); -6.26 to 10.46 | -1.45 (2.98); -6.26 to 9.89 | -1.68 (2.78); -6.26 to 8.78 | -1.83 (2.74); -6.26 to 8.91 | -1.85 (2.74); -6.26 to 9.16 | -1.93 (2.65); -6.26 to 10.46 | -2.09 (2.57); -6.26 to 10.10 | -1.49 (2.95); -6.26 to 10.46 | -1.70 (2.78); -6.26 to 8.91 | -1.79 (2.78); -6.26 to 9.89 | -1.95 (2.67); -6.26 to 8.94 | -1.99 (2.64); -6.26 to 9.41 | -2.03 (2.60); -6.26 to 10.10 |
| Highest education level |  |  |  |  |  |  |  |  |  |  |  |  |  |
| None | 5,224 (8.7%) | 1,872 (10%) | 569 (9.4%) | 499 (8.3%) | 514 (8.5%) | 486 (8.0%) | 1,284 (7.1%) | 1,597 (8.8%) | 440 (7.3%) | 483 (8.0%) | 477 (7.9%) | 1,586 (8.8%) | 641 (11%) |
| Other | 28,887 (48%) | 8,990 (50%) | 2,937 (49%) | 2,962 (49%) | 2,892 (48%) | 2,798 (46%) | 8,308 (46%) | 8,522 (47%) | 2,790 (46%) | 2,828 (47%) | 2,821 (47%) | 8,755 (48%) | 3,171 (53%) |
| University | 26,259 (43%) | 7,199 (40%) | 2,534 (42%) | 2,577 (43%) | 2,615 (43%) | 2,776 (46%) | 8,558 (47%) | 7,972 (44%) | 2,807 (46%) | 2,732 (45%) | 2,752 (45%) | 7,769 (43%) | 2,227 (37%) |
| Physical activity (m<sup>2</sup>/s) | 28.37 (8.07); 4.83 to 69.41 | 26.43 (7.96); 4.83 to 68.08 | 27.94 (7.84); 6.48 to 69.41 | 28.62 (7.84); 9.67 to 68.63 | 28.78 (7.76); 6.46 to 69.21 | 29.20 (7.91); 8.25 to 67.35 | 29.95 (8.07); 7.91 to 68.64 | 28.90 (8.44); 5.69 to 69.41 | 29.27 (8.05); 8.27 to 66.94 | 28.98 (8.04); 4.83 to 66.91 | 28.88 (7.99); 5.12 to 67.23 | 27.89 (7.66); 5.99 to 68.64 | 26.21 (7.80); 5.52 to 69.21 |
| Alcohol (days/wk) | 3.00 (2.50) | 2.85 (2.52) | 2.95 (2.50) | 3.03 (2.51) | 3.01 (2.48) | 3.06 (2.49) | 3.13 (2.49) | 2.98 (2.55) | 3.01 (2.51) | 3.02 (2.50) | 3.03 (2.47) | 3.01 (2.47) | 2.96 (2.48) |
| Smoking |  |  |  |  |  |  |  |  |  |  |  |  |  |
| Never | 34,883 (57%) | 9,611 (53%) | 3,476 (57%) | 3,485 (57%) | 3,540 (58%) | 3,543 (58%) | 11,228 (61%) | 9,793 (54%) | 3,508 (58%) | 3,538 (58%) | 3,559 (58%) | 10,851 (59%) | 3,634 (60%) |
| Previous | 22,148 (36%) | 7,045 (39%) | 2,196 (36%) | 2,230 (37%) | 2,183 (36%) | 2,244 (37%) | 6,250 (34%) | 6,969 (38%) | 2,212 (36%) | 2,155 (35%) | 2,174 (36%) | 6,465 (35%) | 2,173 (36%) |
| Current | 3,805 (6.3%) | 1,577 (8.6%) | 409 (6.7%) | 372 (6.1%) | 353 (5.8%) | 305 (5.0%) | 789 (4.3%) | 1,489 (8.2%) | 357 (5.9%) | 391 (6.4%) | 358 (5.9%) | 935 (5.1%) | 275 (4.5%) |
| Urbanicity |  |  |  |  |  |  |  |  |  |  |  |  |  |
| <10,000 | 9,812 (16%) | 2,660 (15%) | 914 (15%) | 961 (16%) | 1,034 (17%) | 1,033 (17%) | 3,210 (18%) | 2,732 (15%) | 977 (16%) | 936 (16%) | 1,031 (17%) | 3,109 (17%) | 1,027 (17%) |

|  | Overall | Sleep Regularity Index percentiles |  |  |  |  |  | Sleep duration percentiles |  |  |  |  |  |
| --- | --- | --- | --- | --- | --- | --- | --- | --- | --- | --- | --- | --- | --- |
|  | N = 60,998<sup>1</sup> | 0-30<sup>th</sup><br>N = 18,309<sup>1</sup> | 31-40<sup>th</sup><br>N = 6,110<sup>1</sup> | 41-50<sup>th</sup><br>N = 6,083<sup>1</sup> | 51-60<sup>th</sup><br>N = 6,100<sup>1</sup> | 61-70<sup>th</sup><br>N = 6,096<sup>1</sup> | 71-100<sup>th</sup><br>N = 18,300<sup>1</sup> | 0-30<sup>th</sup><br>N = 18,300<sup>1</sup> | 31-40<sup>th</sup><br>N = 6,099<sup>1</sup> | 41-50<sup>th</sup><br>N = 6,100<sup>1</sup> | 51-60<sup>th</sup><br>N = 6,100<sup>1</sup> | 61-90<sup>th</sup><br>N = 18,299<sup>1</sup> | 91-100<sup>th</sup><br>N = 6,100<sup>1</sup> |
| ≥10,000 | 50,599 (84%) | 15,453 (85%) | 5,112 (85%) | 5,080 (84%) | 4,985 (83%) | 5,013 (83%) | 14,956 (82%) | 15,384 (85%) | 5,050 (84%) | 5,102 (84%) | 5,022 (83%) | 15,024 (83%) | 5,017 (83%) |
| Diet |  |  |  |  |  |  |  |  |  |  |  |  |  |
| Healthy | 15,396 (26%) | 4,379 (25%) | 1,422 (24%) | 1,547 (26%) | 1,484 (25%) | 1,564 (26%) | 5,000 (28%) | 4,473 (25%) | 1,532 (26%) | 1,546 (26%) | 1,555 (26%) | 4,734 (27%) | 1,556 (26%) |
| Unhealthy | 43,892 (74%) | 13,334 (75%) | 4,498 (76%) | 4,377 (74%) | 4,444 (75%) | 4,402 (74%) | 12,837 (72%) | 13,272 (75%) | 4,398 (74%) | 4,399 (74%) | 4,405 (74%) | 13,060 (73%) | 4,358 (74%) |
| Sleep Regularity Index | 78.78 (10.69);<br>2.46 to 98.53 | 65.94 (9.57);<br>2.46 to 75.62 | 77.14 (0.85);<br>75.62 to 78.56 | 79.81 (0.71);<br>78.56 to 80.99 | 82.10 (0.63);<br>81.00 to 83.16 | 84.19 (0.60);<br>83.17 to 85.23 | 88.89 (2.56);<br>85.23 to 98.53 | 74.50 (11.91);<br>6.05 to 97.23 | 79.32 (10.00);<br>14.81 to 97.29 | 80.34 (9.64);<br>9.82 to 98.19 | 80.81 (9.33);<br>25.65 to 98.05 | 81.29 (9.25);<br>2.46 to 97.78 | 79.93 (9.90);<br>10.61 to 98.53 |
| Sleep duration (h) | 6.77 (1.00);<br>0.48 to 13.49 | 6.38 (1.23);<br>0.65 to 13.49 | 6.75 (0.94);<br>0.51 to 12.12 | 6.81 (0.89);<br>0.48 to 11.00 | 6.91 (0.86);<br>0.65 to 11.64 | 6.94 (0.83);<br>3.75 to 12.74 | 7.04 (0.76);<br>0.59 to 10.80 | 5.61 (0.71);<br>0.48 to 6.34 | 6.47 (0.07);<br>6.34 to 6.59 | 6.71 (0.07);<br>6.59 to 6.82 | 6.93 (0.06);<br>6.82 to 7.05 | 7.44 (0.25);<br>7.05 to 7.94 | 8.40 (0.46);<br>7.94 to 13.49 |

<sup>1</sup> Mean (SD); Min to Max; n (%)

### Sleep regularity vs. sleep duration across diseases/disorders

Across 199 tested disease and disorder outcomes, irregular sleep (0-30^th^ vs. 71-100^th^ percentiles; SRI < 75.62 vs. SRI ≥ 85.2) predicted significantly higher risk for 131 incident disease/disorder phenotypes (65.8%), whereas short sleep (0-30^th^ vs. 61-90^th^ percentiles; <6.3 h vs. 7.1 to 7.9 h) predicted higher risk for 63 diseases/disorders (31.7%; Model 2 adjustments; FDR<.05; Figures 1–2, Table 2, Supplementary Table S1). Multivariable-adjusted sleep regularity models had superior log-likelihood values compared to equivalent sleep duration models for predicting risk across 115 of the 131 outcomes (87.8%) that were significantly predicted by sleep regularity (and 145 of the total 199 outcomes). Adjustment of significant sleep regularity models for sleep duration improved model fit for 35 of the 131 outcomes (26.7%), based on a likelihood-ratio test. Adjustment of significant sleep duration models for sleep regularity improved model fit for 52 of the 63 outcomes (82.5%).

**Figure 1.**
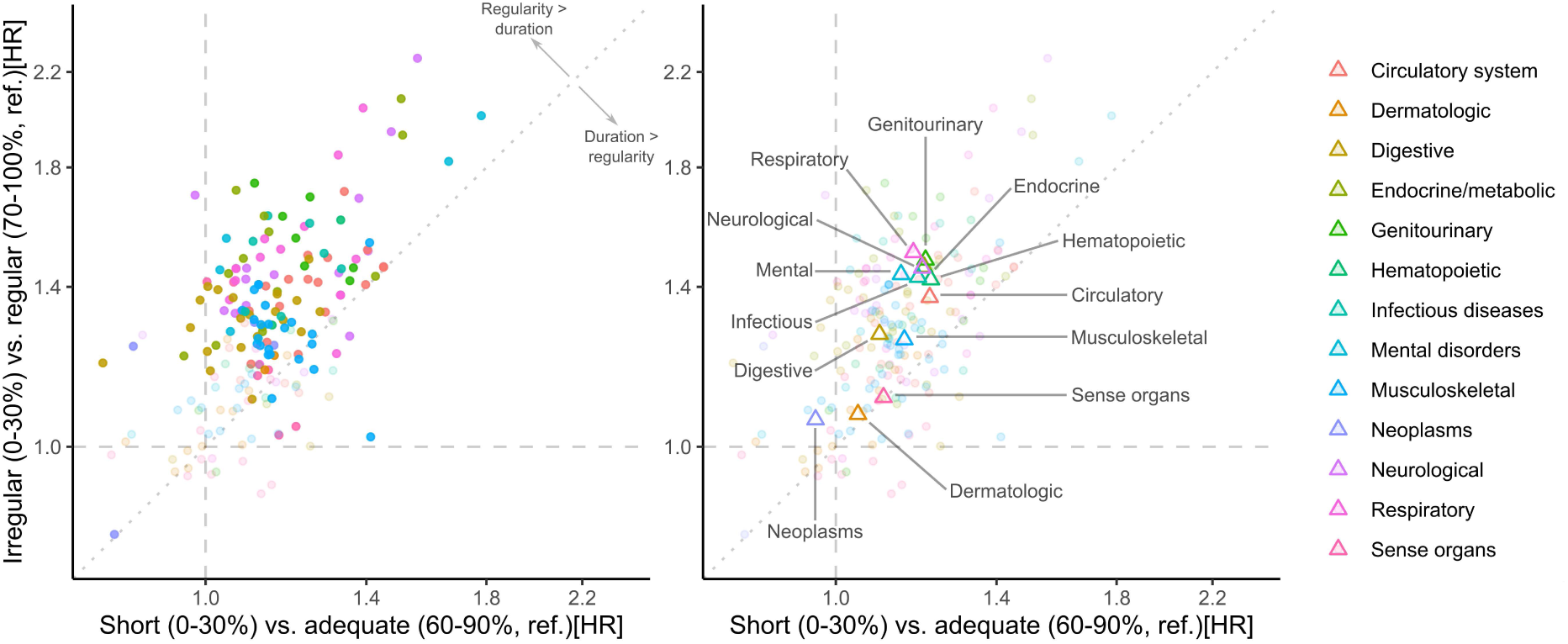
A) Multi-variable adjusted hazard ratio for short vs. adequate (ref.) sleepers (x-axis) and irregular vs. regular (ref.) sleepers (y-axis) as predictors of 199 diseases and disorders. All models were adjusted for age, sex, ethnicity, employment, income, education, and area-level deprivation (Model 2), with sleep regularity and sleep duration in separate models. Opaque point-estimates represent outcomes not significantly associated with sleep regularity or sleep duration (FDR-adjusted p-value <.05). B) Prevalence-weighted centroids of hazard ratios for each of 13 disease and disorder categories.

**Figure 2.**
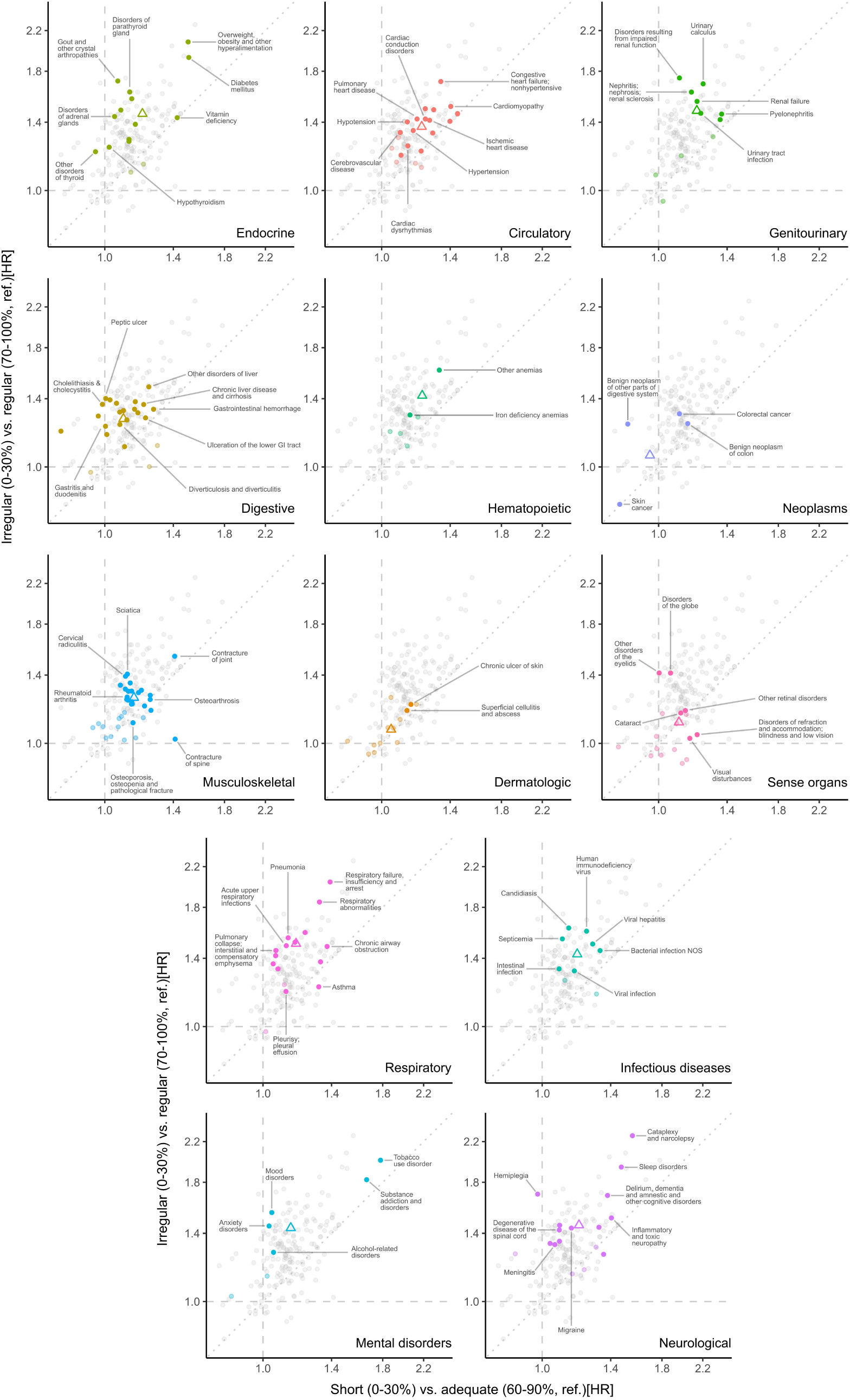
Multi-variable adjusted hazard ratios for short vs. adequate (ref.) sleepers (x-axis) and irregular vs. regular (ref.) sleepers (y-axis) as predictors of 199 diseases/disorders, split according to 13 disease/disorder categories. All models were adjusted for age, sex, ethnicity, employment, income, education, and area-level deprivation (Model 2). Opaque point-estimates represent outcomes not significantly associated with sleep regularity or sleep duration (FDR-adjusted p-value <.05).

**Table 2.** Numbers of diseases/disorders that were significantly predicted by: (i) both irregular sleep and short sleep in separate models; (ii) irregular sleep only; and (iii) short sleep only. Weighted hazard ratios for each outcome category.

|  | Outcomes (N) | Higher risk for irregular and short sleepers (N,%)<sup>a</sup> | Higher risk for irregular sleepers only (N,%) | Higher risk for short sleepers only (N,%) | Weighted hazard ratios |  |
| --- | --- | --- | --- | --- | --- | --- |
|  |  |  |  |  | Irregular (0-30%) vs. regular (70-100%, ref.) [HR] | Short (0-30%) vs. adequate (70-100%, ref.) [HR] |
| Circulatory system | 22 | 13 (59.1) | 4 (18.2) | 0 (0) | 1.37 | 1.22 |
| Dermatologic | 13 | 0 (0) | 2 (15.4) | 0 (0) | 1.07 | 1.05 |
| Digestive | 23 | 6 (30.4) | 13 (56.5) | 0 (0) | 1.27 | 1.10 |
| Endocrine/metabolic | 15 | 6 (40) | 7 (46.7) | 0 (0) | 1.46 | 1.20 |
| Genitourinary | 11 | 5 (45.5) | 2 (18.2) | 0 (0) | 1.48 | 1.21 |
| Hematopoietic | 6 | 1 (16.7) | 1 (16.7) | 0 (0) | 1.42 | 1.22 |
| Infectious diseases | 9 | 2 (22.2) | 5 (55.6) | 0 (0) | 1.43 | 1.19 |
| Mental disorders | 7 | 2 (28.6) | 3 (42.9) | 0 (0) | 1.44 | 1.15 |
| Musculoskeletal | 36 | 10 (27.8) | 12 (33.3) | 2 (5.6) | 1.25 | 1.15 |
| Neoplasms | 4 | 0 (25) | 3 (75) | 0 (0) | 1.06 | 0.96 |
| Neurological | 17 | 6 (35.3) | 7 (41.2) | 0 (0) | 1.46 | 1.20 |
| Respiratory | 18 | 4 (22.2) | 11 (61.1) | 1 (5.6) | 1.51 | 1.18 |
| Sense organs | 18 | 1 (5.6) | 3 (16.7) | 2 (11.1) | 1.11 | 1.10 |
| **Total** | **199** | **56 (28.1)** | **73 (36.7)** | **5 (2.5)** | - | - |

Model estimates were adjusted for age, sex, ethnicity, employment, income, education, and area-level deprivation (Model 2). <sup>a</sup>Each N represents the number of disease/disorder phenotypes that were statistically-significantly predicted by both irregular sleep and short sleep, in separate models (FDR<.05).

Irregular sleep predicted higher risk for 87.0% of digestive outcomes, 86.7% of endocrine/metabolic outcomes, 83.3% of respiratory outcomes, 77.8% of infectious diseases, 77.3% of circulatory outcomes, 76.5% of neurological outcomes, 75.0% of neoplasms, 71.4% of mental disorders, 63.6% of genitourinary outcomes, 61.1% of musculoskeletal outcomes, 33.3% of hematopoietic outcomes, 22.2% of sense organ outcomes, and 15.4% of dermatologic outcomes. By contrast, short sleep predicted higher risk for 59.1% of circulatory outcomes, 45.5% of genitourinary outcomes, 40.0% of endocrine/metabolic outcomes, 35.3% of neurological outcomes, 31.7% of musculoskeletal outcomes, 30.4% of digestive outcomes, 28.6% of mental disorders, 27.8% of respiratory outcomes, 22.2% of infectious diseases, 16.7% of sense organ outcomes, and 16.7% of hematopoietic outcomes.

Of 131 models with significant regularity-outcome associations, 90 (68.7%) of these had superior log-likelihood values, and therefore a better model fit, compared with equivalent duration-only models and were also not improved by adjustment for sleep duration. By contrast, 63 models with significant duration-outcome associations, 9 (14.3%) had superior log-likelihood values compared with equivalent SRI-only models and were also not improved by adjustment for SRI. Outcomes most strongly predicted by regularity and duration are listed in Table 3, ranked by hazard ratio. Across the top 30 outcomes, the most represented categories for regularity were endocrine/metabolic (N=5), neurological (N=5), respiratory (N=5), genitourinary (N=4), infectious diseases (N=4), and mental disorders (N=3). By contrast, the most represented categories for duration were circulatory system (N=7), neurological (N=6), respiratory (N=4), and endocrine/metabolic (N=3).

**Table 3.**
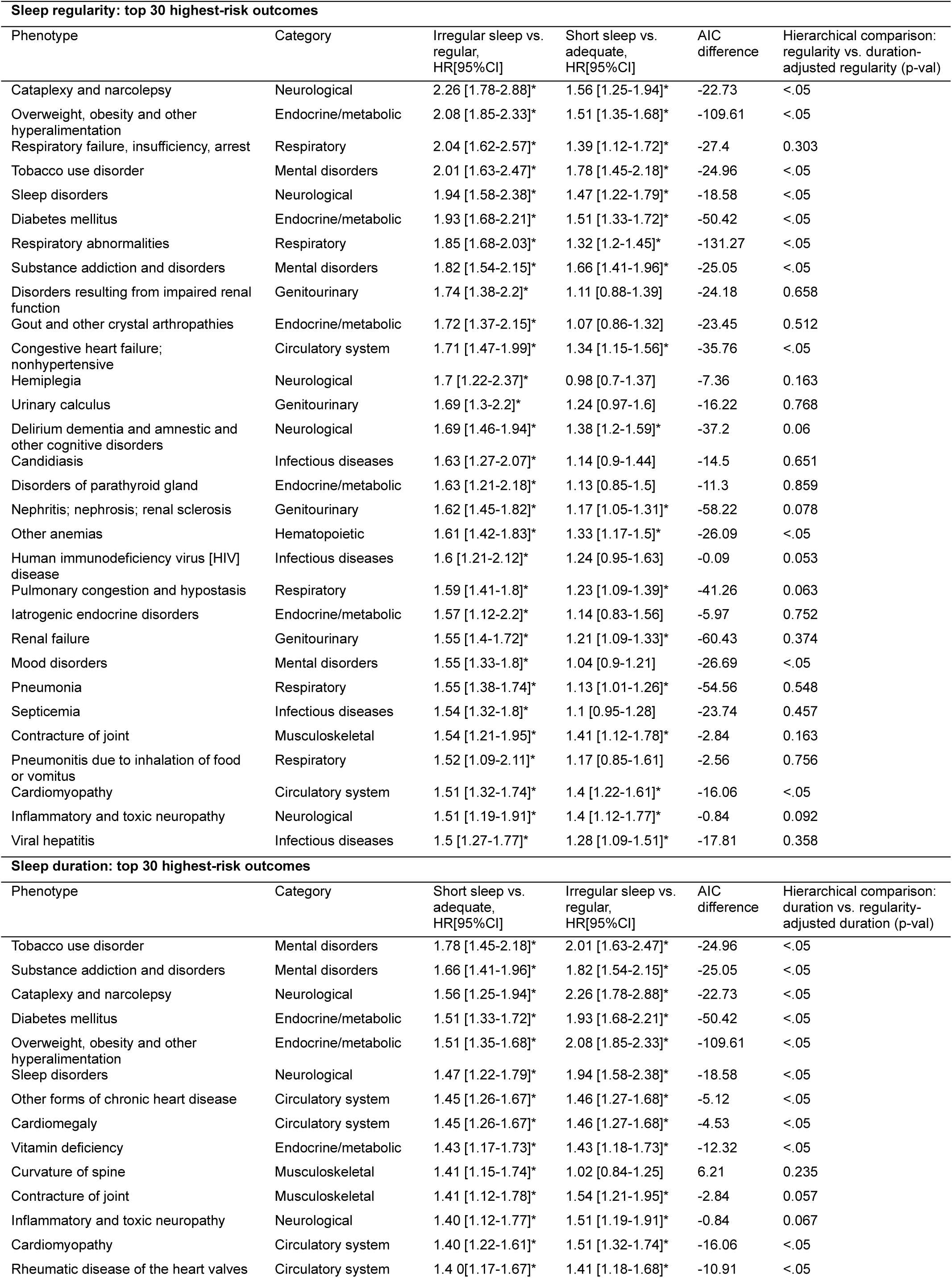
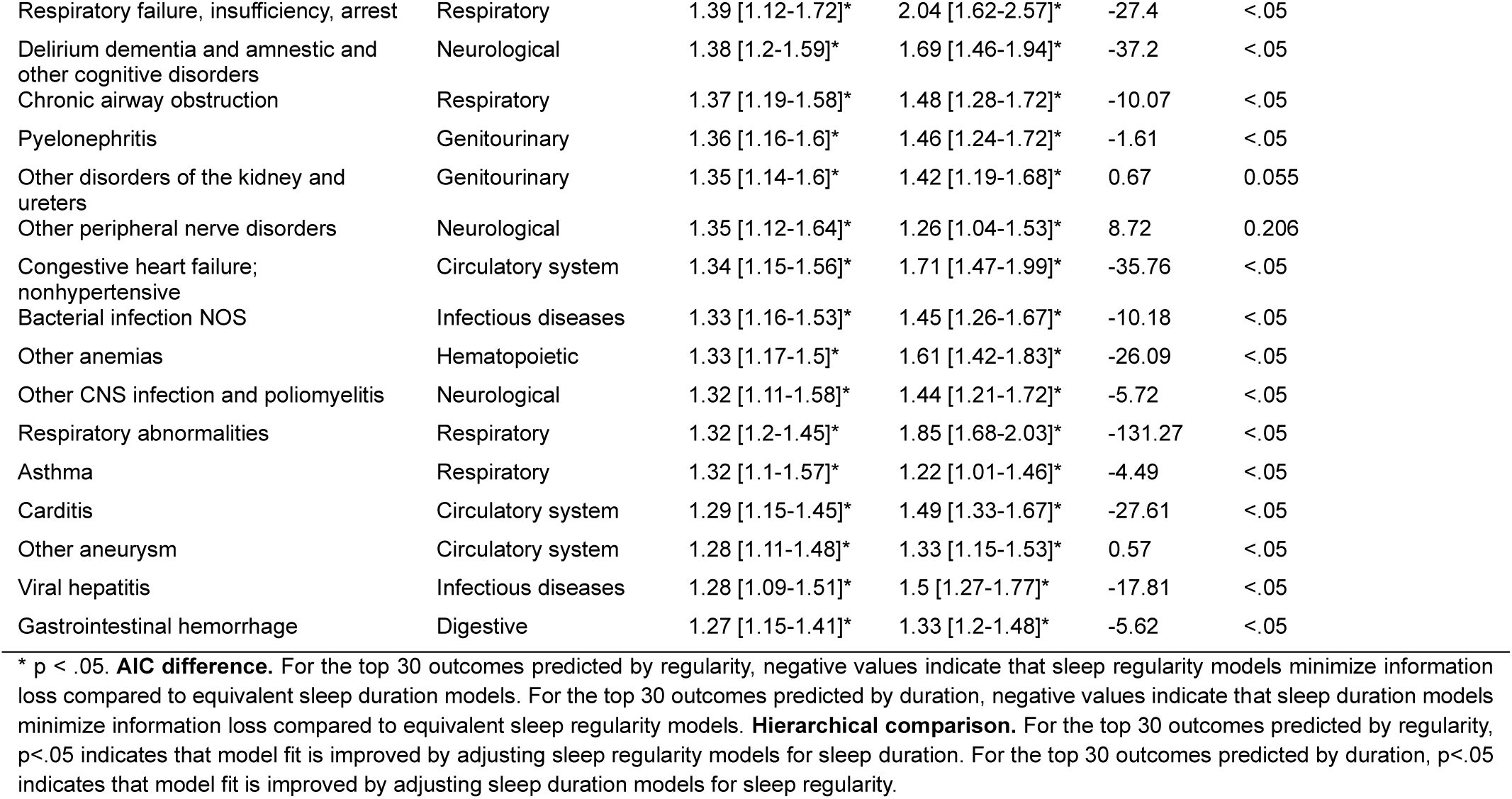
Top 30 highest-risk disease and disorder outcomes associated with irregular sleep and/or short sleep duration, ordered by hazard ratio, and including regularity vs. duration model comparisons.

### Sex differences

Across the top 30 incident diseases/disorders (Table 4), the categories most consistently linked to irregular sleep among men were Respiratory (N=11), Neurological (N=5), Endocrine/metabolic (N=3), Infectious (N=3), and Genitourinary (N=3), whereas these categories among women were Circulatory (N=8), Respiratory (N=6), Endocrine/metabolic (N=5), Neurological (N=3), and Mental disorders (N=3). By contrast, the categories most consistently linked to short sleep among men were Circulatory (N=7), Musculoskeletal (N=7), Neurological (N=3), Endocrine/metabolic (N=3), and Genitourinary (N=3), whereas these categories among women were Circulatory (N=8), Respiratory (N=4), Endocrine/metabolic (N=4), and Digestive (N=4). A complete list of regularity and duration as predictors of risk for incident diseases/disorders, stratified by sex, is provided in Supplementary Table S2.

**Table 4.**
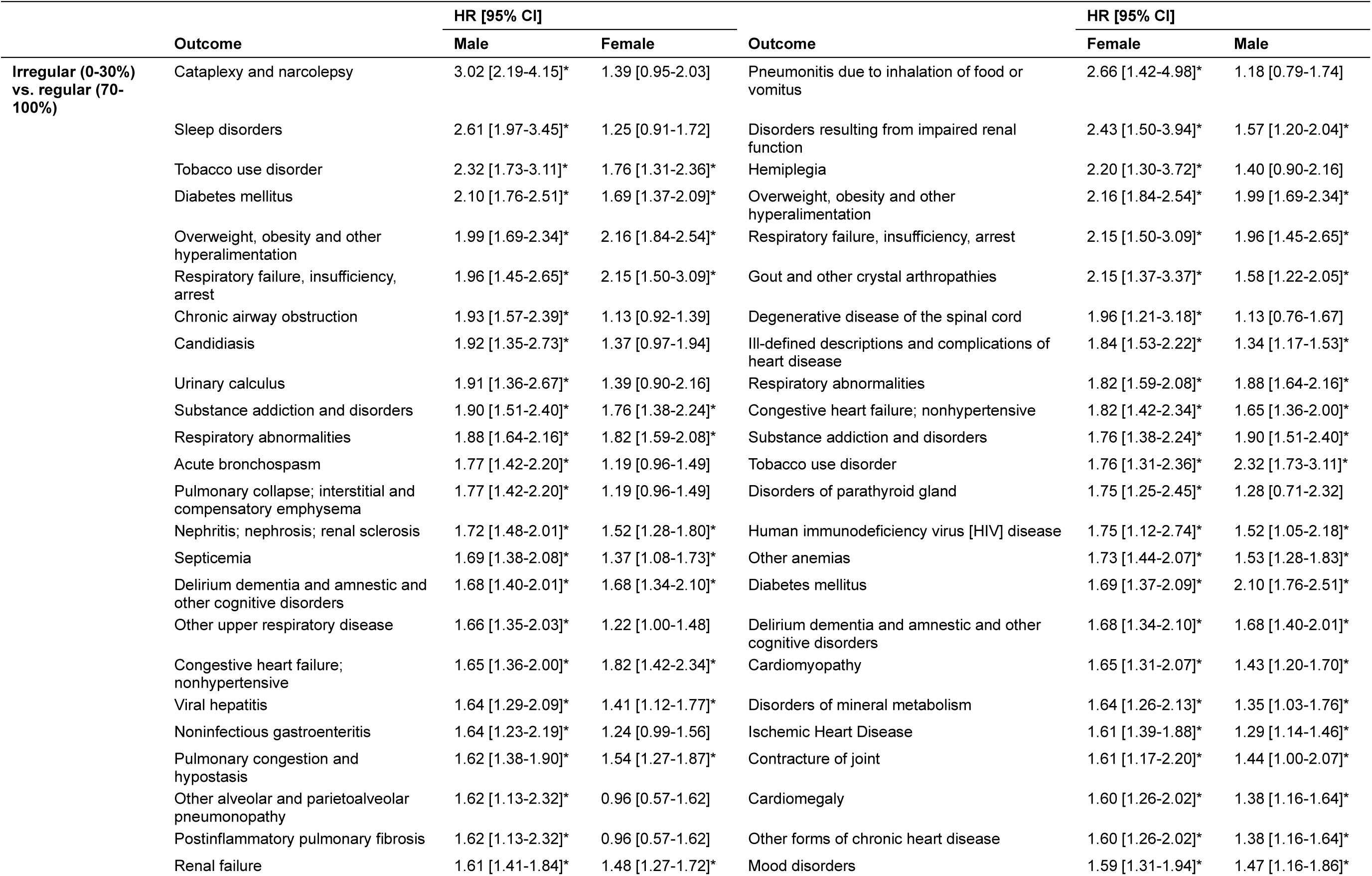
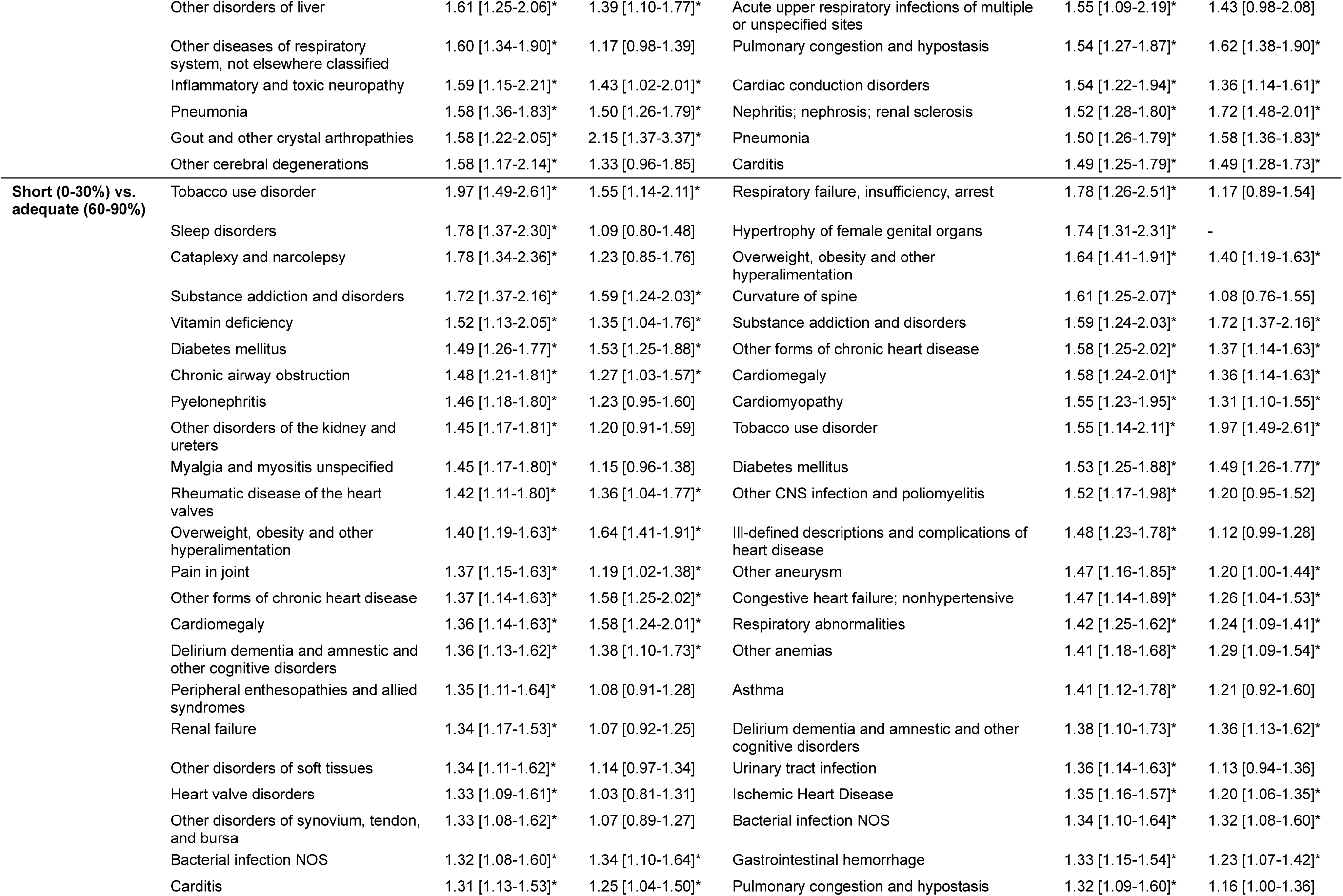
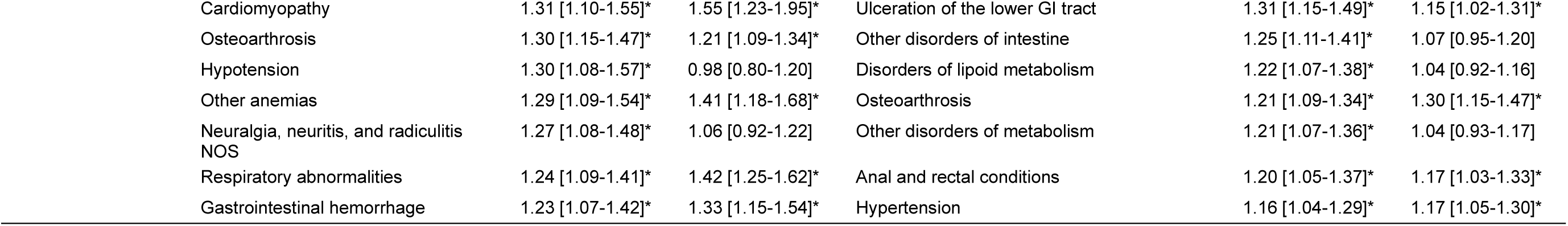
Top 30 disease and disorder outcomes associated with irregular and/or short sleep, for males and females, ordered by hazard ratio.

## Discussion

Across 199 incident outcomes spanning the human disease/disorder phenome, we found that sleep regularity was a more robust predictor of risk for disease/disorder incidence than sleep duration. Across the 199 disease/disorder outcomes, regularity was a superior predictor to duration of risk for 90 outcomes, while duration was a superior predictor to regularity for only 9 outcomes, based on comparison of both independent and hierarchical models. We also found notable sex differences in sleep regularity and sleep duration as predictors of risk for diseases/disorders, highlighting the need for greater consideration of sex when developing sleep or circadian health interventions.

Metabolic and endocrine conditions were among the strongest outcomes with elevated risk in the models reported. Of the top 10 diseases predicted by both sleep regularity and sleep duration, three were metabolic/endocrine. Sleep regularity and duration are both well-established risk factors for type 2 diabetes and obesity/overweight in observational studies.^13,14,24,39–42^ Experimental studies have demonstrated negative effects of sleep restriction on metabolic function,^43^ but also reveal circadian disruption as a causal mechanism for metabolic dysregulation, independent from sleep duration.^44–48^ Here, we found that irregular sleep was associated with 93% and 108% higher risk for diabetes and obesity/overweight, respectively, while short sleep was associated with 51% higher risk for both outcomes. These findings point to the necessity of maintaining both regular and adequate sleep to reduce metabolic risk.

Irregular and short sleep were both robust risk factors for circulatory system outcomes in this cohort, consistent with observational and experimental literature. Previous observations show higher risks of major adverse cardiovascular events and cardiovascular diseases for people with sleep duration outside 6-9h^49,50^ and with irregular sleep timing.^13,15,51^ Experimental disruption of light-dark and sleep-wake patterns causes elevated blood pressure and heart rate, inflammation, and hypercoagulability,^44,52–54^ likely driven by disruption of cardiovascular circadian rhythms.^55–61^ Long-term circadian disruption causes hypertrophy, myocardial fibrosis, impaired contractility, and progression to heart failure in animal studies.^62–64^ In this cohort, irregular sleepers had 25-71% higher risks of heart failure, ischemic heart disease, hypertension, cardiac dysrhythmias, cerebrovascular diseases, pulmonary heart disease, and cardiomyopathy, whereas short sleepers had 10-40% higher risks of these outcomes. Higher risks for irregular *vs*. short sleepers are consistent with previous reports in smaller samples.^13,65^ Sex differences in circulatory system risks were observed: in the top 30 outcomes for men, risk for one outcome was predicted by irregular sleep vs. six outcomes for short sleep, whereas women had risk for six and seven outcomes predicted by irregular and short sleep, respectively. These findings are consistent with previous observations of higher cardiovascular risks for women who are shift workers and/or who have brighter nights,^37,66^ and with greater circadian sensitivity to light in women.^67^

Risk for respiratory diseases was prospectively predicted by sleep, with generally stronger associations for irregular sleep. We found irregular sleep was a stronger predictor than short sleep for risk of new ‘respiratory failure, insufficiency and arrest’ (higher risk of 104% vs. 39%, respectively); pneumonia (55% vs. 13%); and acute upper respiratory infections (49% vs.12%). Higher risks for irregular and short sleepers were observed for chronic airway obstruction (48% vs. 37%) and asthma (22% vs. 32%), respectively. Notably, women showed a stronger association of pneumonitis due to food/vomit inhalation with irregular sleep (166% increased risk for women vs. *n.s.* for men) and ‘respiratory failure / insufficiency / arrest’ with short sleep (78% vs. *n.s.*). Meanwhile, men showed a stronger association of ‘pulmonary collapse; interstitial and compensatory emphysema’ with irregular sleep (77% for men vs. *n.s.* for women) and chronic airway obstruction (e.g., COPD) with short sleep (48% vs. 27%), suggesting significant sex-based differences in sleep-related vulnerability to respiratory diseases. Some prior research indicates irregular sleepers and shift workers have higher risk of respiratory outcomes.^68,69^ We note the possibility of reverse causation in our sample, since sleep disruption, particularly due to nocturnal apnea/hypopnea, is common in many respiratory diseases.^70–72^ Although we applied a 1-year exclusionary window for diagnoses after sleep assessment, some respiratory diseases are likely to have been undiagnosed in some individuals for longer than this. Circadian rhythms in respiratory function^73^ have been observed for airway caliber,^74^ ventilatory response to hypercapnia,^75^ forced expiratory volume and airway resistance,^76^, and both oxygen desaturation and apnea/hypopnea during sleep.^77^ Furthermore, the asthmatic response to allergens exhibits a diurnal rhythm.^78,79^ Irregular sleep patterns, which disrupt circadian rhythms, are therefore a plausible causal risk factor for developing respiratory diseases. Short sleep duration may contribute to risk via decreased immune function and increased risk of acquiring upper airway tract infections,^80^ which can exacerbate respiratory diseases.

Renal disorders and sleep share bidirectional links.^81^ Sleep disturbances are common in renal patients,^82^ and short sleepers are at higher risk of chronic kidney disease.^83^ However, links between sleep regularity and kidney diseases remain mostly unexplored.^84^ Here, irregular sleep was a stronger predictor than short sleep for increased risk of nephritis/ nephrosis/ renal sclerosis’: 62% (irregular) vs. 17% (short); renal failure (55% vs. 21%); kidney stones (69% vs. 24%); and disorders resulting from impaired renal function (74% vs. 11%). Similarly, higher risks for irregular *vs*. short sleepers were observed for urinary tract infections (46% vs. 23%), and pyelonephritis (46% vs. 36%). Circadian kidney rhythms^85^ are present in renal water excretion, glomerular filtration rate, and renal plasma flow,^86^ and regular physical activity rhythms are a suggested entrainment mechanism for circadian kidney rhythms.^87^ Irregular sleep has been linked with hypertension^88^ and diabetes,^14^ which are risk factors for kidney dysfunction and may contribute to the observed renal outcomes. Irregular sleepers also had an increased risk of ‘Other anemias’ (including ‘anemia in chronic kidney disease’) compared to short sleepers (61% vs. 33%). This could be explained by shifted or blunted rhythms in renal erythropoietin release,^89^ which stimulates red-blood cell production (anemia is common in chronic kidney disease^90^), or by disruption of rhythms in bone marrow cell proliferation.^91^ We observed sex differences in the associations of irregular sleep with renal-related outcomes. For women, higher comparative risks were observed for disorders resulting from impaired renal function (including renal osteodystrophy; 143% for women vs. 57% for men) and gout (115% vs. 58%), whereas men had higher comparative risk than women for kidney stones (91% for men vs. 39% for women). These findings are consistent with sex differences in clinical presentation of hyperparathyroidism^92^ (a downstream complication of kidney disease), and the coupling of central circadian rhythms with estrous cyclicity.^93^

Irregular sleep demonstrated consistently strong associations with risk for musculoskeletal outcomes, often exceeding those observed for short sleep duration, including sciatica (41% vs. 12% higher risk), cervical radiculitis (39% vs. 11%), rheumatoid arthritis (26% vs. 12%), and spinal curvature (41% vs. 2%), while ‘osteoporosis / osteopenia / pathological fracture’ and osteoarthrosis showed similar associations for both sleep dimensions. These findings align with evidence that musculoskeletal tissues are regulated by circadian rhythms controlling bone,^94,95^ muscle,^96,97^ cartilage,^98^ and inflammatory processes.^99,100^ Experimentally induced short and irregular sleep impairs bone formation in humans,^101^ while animal studies demonstrate that circadian disruption contributes to intervertebral disc degeneration,^102,103^ providing a potential mechanistic explanation for the particularly strong associations with disc-related disorders such as sciatica and cervical radiculitis. Irregular sleep may also reflect irregular rest-activity and biomechanical loading patterns^104^ that contribute to degeneration and impaired tissue repair. Circadian disruption alters hormonal pathways involved in musculoskeletal maintenance, for example melatonin-mediated bone formation.^105^ However, substantial bidirectionality is likely, as chronic pain, immobility, and difficulty finding comfortable sleeping positions may disrupt sleep, and substantial diagnostic delays can occur for musculoskeletal conditions.^106,107^ Overall, the stronger and more consistent associations with sleep irregularity suggest circadian disruption plays an important role in musculoskeletal degeneration.

Disruption of sleep and circadian rhythms is a hallmark of most mental health conditions.^108–111^ Consistent with this, we found sleep regularity predicted risk for 71%, and duration predicted risk for 29%, of the mental health conditions examined. The strongest associations were found for substance use disorder, tobacco use disorder, and mood disorders. Sleep disruptions can both arise from these conditions and contribute to their ongoing nature and severity,^112,113^ meaning these associations likely reflect bidirectional causal relationships. Indeed, recent research has found that individuals with alcohol use disorder are at greater risk of relapse if they have lower SRI.^114^ In the UK Biobank cohort, we found important sex-specific differences in these associations. Specifically, addictive conditions were more strongly associated with both sleep regularity and duration in men, whereas in women mood disorders were more strongly associated with sleep regularity and duration. These findings suggest that the effect of sleep-targeted interventions to improve mental health may differ according to sex, and the usage of sleep regularity or duration as predictive markers of mental health needs to be contextualized differently for men and women.

Among neurological conditions, sleep regularity and duration were strong predictors of risk for delirium, dementia and amnestic and other cognitive disorders (67% vs. 38% higher risks for irregular vs. short sleepers). These findings are consistent with dose-response analyses for sleep duration and regularity with incident all-cause dementia previously published in the same dataset.^115^ Disrupted sleep and circadian rhythms are not only prominent symptoms, but also potential drivers of neurodegeneration.^116^ However, evidence for short sleep duration as a risk modifier for dementia is mixed based on meta-analyses of prospective studies.^117^ We also found that sleep regularity predicted a 44% higher risk of migraine, whereas sleep duration was not associated with migraine. Consistent with this, previous research has found that low sleep efficiency (which may be coincident with irregular sleep through fragmentation of the sleep episodes) is associated with migraine occurrence on the following day, whereas short sleep duration is not.^118^ These findings suggest sleep regularity may be a promising target for managing or mitigating neurological outcomes.

Unsurprisingly, both sleep regularity and duration were strong predictors of risk for sleep disorders (a classification which includes sleep apnea, insomnia, circadian rhythm sleep disorders, and parasomnias) and cataplexy/narcolepsy. Many of these disorders manifest as fragmented or irregular sleep timing,^68,119^ or reduced sleep duration. Surprisingly, however, these associations were only significant in men. This may reflect differing prevalences, clinical presentation, and/or severity of sleep disorders in this cohort: sleep apnea is more common in men,^120^ while insomnia is more common in women.^121^ We also note that sleep disorders may be underdiagnosed in the UK Biobank.^122^

Irregular sleepers had consistently stronger risk of infectious diseases than short sleepers, including viral infection (32% vs. 17% higher risk), candidiasis (63% vs. 14%), intestinal infection (33% vs. 9%), HIV/AIDS (60% vs. 24%), septicemia (54% vs. 10%), and viral hepatitis (50% vs. 28%). Mechanistic evidence demonstrates that circadian disruption impairs immune function, independent of sleep deprivation.^123^ Circadian rhythms regulate leukocyte trafficking, including release from bone marrow, homing to organs, recruitment to infected tissues, phagocytic clearance, and lymphatic exit, through coordinated neuroimmune and endocrine signaling pathways.^124^ Cortisol and catecholamine rhythms also shape daily rhythms in immune cell populations, with naive T cells reduced during the active phase and effector cells peaking during the day.^125^ Experimental studies suggest that circadian disruption may impair antiviral defense mechanisms through altered interferon signaling and clock gene function, including PER2-mediated regulation of interferon-γ responses and reciprocal effects of interferon signaling on central clock function.^126–129^ Together, these mechanisms provide biological plausibility for the strong associations observed with sleep regularity.

We did not find significant associations of short sleep or irregular sleep with most cancer outcomes. Irregular sleep was associated with a 30% higher risk of colorectal cancer, with no significant association for short sleep. This finding is consistent with other large-scale studies finding no association between short sleep duration and colorectal cancer,^130^ whereas a metric that combines non-optimal duration with other aspects of sleep disturbance such as insomnia, snoring, and daytime sleepiness predicted higher colorectal cancer risks.^131^ Skin cancer was unique among all 199 diseases examined in showing reduced risks for both short sleep (-17%) and irregular sleep (-17%). This finding may be explained by individuals with unhealthy sleep patterns having reduced time outdoors, mitigating skin cancer risk.

There are several important limitations. First, reverse-causality in the relationships of sleep with disease/disorder outcomes is possible, despite the longitudinal study design and our follow-up period commencing 1 year after activity-tracking. Those with sub-clinical or non-diagnosed outcomes could have short and/or irregular sleep patterns for years prior to their diagnosis, since diagnostic delay is common for many conditions.^106,107,132–134^ Second, while we accounted for healthy diet, we note that meal timing was not directly measured in this dataset. Irregular sleep is likely linked with irregular meal timing, which could mediate observed relationships of sleep regularity with some diseases/disorders, via disruption of peripheral circadian clocks (*e.g*., liver, pancreas).^135^ Third, the UK Biobank population is not representative of the global population (*e.g*., ethnicity, sex, socioeconomic status, lifestyle, and health status).^136^ Finally, comorbidity and multi-morbidity were outside the broad scope of the present study, but require investigation: for many conditions, it is unclear whether sleep itself, or the prior existence of a sleep-related disease/disorder, is the key proximal risk factor (*e.g*., irregular sleep may play a causal role in developing type 2 diabetes, but type 2 diabetes is also a risk factor for subsequent cardiovascular disease).

Our findings demonstrate that sleep regularity is a stronger prospective predictor of risk for human disease/disorder than sleep duration across major physiological systems in this cohort, suggesting regular sleep is a fundamental component of healthy physiological and psychological functioning. These findings are consistent with other large-scale phenome-wide association studies, which demonstrate that variable sleep duration predicts a larger number of diseases/disorders than inadequate sleep duration,^137^ and that sleep regularity is a key feature of multi-dimensional sleep metrics that predict risk for a broad spectrum of diseases.^138^ Here, we found that irregular sleep predicted 13 of the top 25 health outcomes with the greatest global burden,^139^ based on disability-adjusted life years, including: ischemic heart disease, stroke, lower respiratory infections, COPD, diabetes, low back pain, depressive disorders, headache disorders, cirrhosis, musculoskeletal disorders, chronic kidney disease, anxiety disorders, and HIV/AIDS. Given the pervasive role of circadian rhythms in regulating physiology,^21^ circadian disruption likely underpins the broad and robust associations observed. These findings continue to challenge sleep duration as the most important dimension of sleep health, and indicate that improving sleep regularity should be explored as a strategy for preventing disease and promoting long-term health.

## Data Availability

All data produced are available online via application to the UK Biobank.

## Supplementary

**Table S1.** Risk estimates across disease/disorder outcomes for irregular and short sleepers, including sleep regularity vs. sleep duration model comparisons, adjusted across four hierarchical model levels.

**Table S2.** Risk estimates across disease/disorder outcomes for irregular and short sleepers, split by male and female, adjusted across four hierarchical model levels.

**Table S3.** STROBE Checklist

## Notes

### Competing Interest Statement

B. L. has received research grants from Withings, OURA, Medical Research Future Fund and NHMRC. A.C.R. received research funding from the Australian Research Council (Discovery Early Career Researcher Award - DE250101060). M.K.R. has consulted for Eli Lilly and has modest stock ownership in GSK. M.K.R. disclosures are not related to the current work. F.A.J.L.S. served on the Board of Directors for the Sleep Research Society and has consulted for the University of Alabama at Birmingham and Morehouse School of Medicine. F.A.J.L.S. interests were reviewed and managed by Brigham and Women's Hospital and Partners HealthCare under their conflict-of-interest policies. F.A.J.L.S. consultancies are not related to the current work. R.J.A. has research funding/support from the National Health and Medical Research Council, the Medical Research Future Fund, the Hospital Research Foundation, Flinders Foundation, Sydney Trains, Safework SA, Withings, ResMed Foundation, Neuroflex, Philips and the Lifetime Support Authority for research related to sleep. S.W.C. has consulted for Dyson. A.J.K.P. and S.W.C. are co-founders and co-directors of Circadian Health Innovations PTY LTD. A.J.K.P. and S.W.C. have received research funding from Versalux and Delos. S.W.C. has received research funding from Beacon Lighting. D.P.W., A.C.B., H.S., D.S., K.S., and R.S. have no conflicts of interest to disclose.

### Author Declarations

This research was conducted using UK Biobank data (Project ID: 6818), and ethical approval was granted by the North West Multi-centre Research Ethics Committee.

