## Supplementary Table S1 for "Sleep regularity outweighs sleep duration as a predictor of disease": Supplementary Table S1.html

<!DOCTYPE html>


\* FDR-adjusted p < .05. **AIC difference:** Negative values indicate that sleep regularity models
minimize information loss compared to equivalent sleep duration models.
**Hierarchical comparison:** p<.05 indicates
that model fit is improved by adjusting sleep regularity models
for sleep duration, or by adjusting sleep duration models for sleep regularity
