## Supplementary Table S3 for "Sleep regularity outweighs sleep duration as a predictor of disease"

**Table S3. STROBE Checklist**

STROBE Statement—Checklist of items that should be included in reports of ***cohort studies***

|  | **Item No** | **Recommendation** | **Manuscript Section** |
| --- | --- | --- | --- |
| **Title and abstract** | 1 | (*a*) Indicate the study’s design with a commonly used term in the title or the abstract | - Abstract |
|  |  | (*b*) Provide in the abstract an informative and balanced summary of what was done and what was found | - Abstract |
| **Introduction** | | |  |
| Background/rationale | 2 | Explain the scientific background and rationale for the investigation being reported | - Introduction |
| Objectives | 3 | State specific objectives, including any prespecified hypotheses | - Abstract, Introduction, Methods |
| **Methods** | | |  |
| Study design | 4 | Present key elements of study design early in the paper | - Title, Abstract, Introduction, Methods |
| Setting | 5 | Describe the setting, locations, and relevant dates, including periods of recruitment, exposure, follow-up, and data collection | - Methods |
| Participants | 6 | (*a*) Give the eligibility criteria, and the sources and methods of selection of participants. Describe methods of follow-up | - Methods |
|  |  | (*b*) For matched studies, give matching criteria and number of exposed and unexposed |  |
| Variables | 7 | Clearly define all outcomes, exposures, predictors, potential confounders, and effect modifiers. Give diagnostic criteria, if applicable | - Abstract, Introduction, Methods |
| Data sources/ measurement | 8* | For each variable of interest, give sources of data and details of methods of assessment (measurement). Describe comparability of assessment methods if there is more than one group | - Methods |
| Bias | 9 | Describe any efforts to address potential sources of bias | - Methods |
| Study size | 10 | Explain how the study size was arrived at | - Methods |
| Quantitative variables | 11 | Explain how quantitative variables were handled in the analyses. If applicable, describe which groupings were chosen and why | - Methods |
| Statistical methods | 12 | (*a*) Describe all statistical methods, including those used to control for confounding | - Methods |
|  |  | (*b*) Describe any methods used to examine subgroups and interactions | - Methods |
|  |  | (*c*) Explain how missing data were addressed | - Methods |
|  |  | (*d*) If applicable, explain how loss to follow-up was addressed | - Methods |
|  |  | (*e*) Describe any sensitivity analyses | - Methods, Supplement |
| **Results** | | |  |
| Participants | 13* | (a) Report numbers of individuals at each stage of study—eg numbers potentially eligible, examined for eligibility, confirmed eligible, included in the study, completing follow-up, and analysed | - Methods |
|  |  | (b) Give reasons for non-participation at each stage | - Methods |
|  |  | (c) Consider use of a flow diagram |  |
| Descriptive data | 14* | (a) Give characteristics of study participants (eg demographic, clinical, social) and information on exposures and potential confounders | - Results |
|  |  | (b) Indicate number of participants with missing data for each variable of interest | - Results |
|  |  | (c) Summarise follow-up time (eg, average and total amount) | - Results |
| Outcome data | 15* | Report numbers of outcome events or summary measures over time | - Supplement |
| Main results | 16 | (*a*) Give unadjusted estimates and, if applicable, confounder-adjusted estimates and their precision (eg, 95% confidence interval). Make clear which confounders were adjusted for and why they were included | - Results, Supplement |
|  |  | (*b*) Report category boundaries when continuous variables were categorized | - Methods, Results |
|  |  | (*c*) If relevant, consider translating estimates of relative risk into absolute risk for a meaningful time period |  |
| Other analyses | 17 | Report other analyses done—eg analyses of subgroups and interactions, and sensitivity analyses | - Supplement |
| **Discussion** | | |  |
| Key results | 18 | Summarise key results with reference to study objectives | - Discussion |
| Limitations | 19 | Discuss limitations of the study, taking into account sources of potential bias or imprecision. Discuss both direction and magnitude of any potential bias | - Discussion |
| Interpretation | 20 | Give a cautious overall interpretation of results considering objectives, limitations, multiplicity of analyses, results from similar studies, and other relevant evidence | - Discussion |
| Generalisability | 21 | Discuss the generalisability (external validity) of the study results | - Discussion |
| **Other information** | | |  |
| Funding | 22 | Give the source of funding and the role of the funders for the present study and, if applicable, for the original study on which the present article is based |  |
